## Supplemental Data for "Dose response of running on blood biomarkers of wellness in the generally healthy"

**Table 1S**. Number of people in each category by age group. Significant trend toward younger individuals reporting higher running volume, with more than 75% of the elite group falling between the ages of 18 and 35.

| Age Group | PRO | HVAM | MVAM | LVAM | SED |
|---|---|---|---|---|---|
| 18-35 | 53 | 434 | 1975 | 3452 | 1032 |
| 35-45 | 25 | 366 | 2537 | 3964 | 1431 |
| 45-55 | 4 | 218 | 1606 | 2469 | 1152 |
| >55 | NA | 85 | 629 | 992 | 813 |

### Table 2S. Full running volume vs. blood biomarker results

| Biomarker | ANOVA p-value | Trend p-value | lowest mean | highest mean |
|---|---|---|---|---|
| Alb | <1e-16 | <0.001 | MVAM | PRO |
| ALT | <1e-16 | <1e-16 | SED | PRO |
| AST | <1e-16 | <0.001 | SED | PRO |
| B12 | <0.001 | <0.001 | SED | PRO |
| BASOS | 0.001 | 0.004 | LVAM | PRO |
| BASOS_PCT | <0.001 | 0.156 | SED | PRO |
| Ca | 0.007 | 0.030 | MVAM | PRO |
| Chol | <0.001 | 0.005 | PRO | SED |
| CK | <1e-16 | <1e-16 | SED | PRO |
| Cor | <0.001 | 0.675 | SED | PRO |
| D | <1e-16 | 0.424 | SED | PRO |
| DHEAS | <0.001 | <0.001 | SED | PRO |
| EOS | <0.001 | 0.371 | HVAM | SED |
| EOS_PCT | <0.001 | 0.137 | HVAM | MVAM |
| FE | <0.001 | 0.119 | SED | PRO |
| Fer | <1e-16 | <1e-16 | MVAM | SED |
| Fol | <1e-16 | <0.001 | SED | PRO |
| FT | <0.001 | 0.013 | SED | PRO |
| GGT | <1e-16 | <0.001 | PRO | SED |
| Glu | 0.087 | 0.184 | PRO | SED |
| Hb | 0.002 | <0.001 | MVAM | PRO |
| HCT | 0.053 | 0.055 | MVAM | PRO |
| HDL | <1e-16 | <0.001 | SED | PRO |
| HbA1c | <0.001 | 0.010 | PRO | SED |
| hsCRP | <0.001 | 0.176 | PRO | SED |
| K | <1e-16 | <0.001 | SED | LVAM |
| LDL | <0.001 | 0.006 | PRO | SED |
| LYMPHS | <0.001 | 0.008 | PRO | SED |
| LYMPHS_PCT | <1e-16 | 0.417 | SED | PRO |
| MCH | 0.197 | 0.077 | SED | PRO |
| MCHC | <1e-16 | 0.276 | SED | PRO |
| MCV | <0.001 | <0.001 | SED | PRO |
| Mg | <0.001 | 0.276 | PRO | SED |
| MONOS | <0.001 | 0.175 | PRO | SED |
| MONOS_PCT | <0.001 | 0.137 | SED | LVAM |
| MPV | 0.058 | 0.089 | SED | HVAM |
| Na | <1e-16 | 0.622 | HVAM | SED |
| NEUT | <0.001 | 0.007 | PRO | SED |
| NEUT_PCT | <0.001 | 0.764 | PRO | SED |
| PLT | <0.001 | 0.058 | LVAM | SED |
| RBC | 0.016 | 0.880 | MVAM | SED |
| RBC_Mg | <0.001 | 0.773 | PRO | SED |
| RDW | <1e-16 | 0.002 | PRO | SED |
| SHBG | <1e-16 | 0.004 | SED | PRO |
| Tes | <1e-16 | 0.675 | MVAM | LVAM |
| Tg | <1e-16 | <1e-16 | PRO | SED |
| TIBC | <0.001 | 0.417 | LVAM | MVAM |
| TS | <1e-16 | 0.298 | SED | PRO |
| WBC | <1e-16 | <1e-16 | PRO | SED |

**Table 3S. 2S-MR results with BMI as the exposure and select biomarkers as outcomes**

| Exposure-outcome | Method | Causal estimate | | | |
|---|---|---|---|---|---|
| | | SNP | Beta | SE | *p* |
| BMI-hsCRP | MR Egger | 233 | 0.440 | 0.054 | 1.74E-14 |
| | Weighted median | 233 | 0.385 | 0.022 | 7.56E-71 |
| | Inverse variance weighted | 233 | 0.352 | 0.018 | 6.78E-84 |
| | Simple mode | 233 | 0.403 | 0.067 | 8.11E-09 |
| | Weighted mode | 233 | 0.413 | 0.039 | 9.98E-22 |
| Test for horizontal pleiotropy: MR Egger intercept = -0.00191, SE = 0.0109, p = 0.0816 | | | | | |
| BMI-HbA1c | MR Egger | 213 | 0.084 | 0.042 | 4.57E-02 |
| | Weighted median | 213 | 0.036 | 0.022 | 9.98E-02 |
| | Inverse variance weighted | 213 | 0.031 | 0.014 | 3.16E-02 |
| | Simple mode | 213 | 0.021 | 0.055 | 7.07E-01 |
| | Weighted mode | 213 | 0.080 | 0.036 | 2.88E-02 |
| Test for horizontal pleiotropy: MR Egger intercept = -0.00115, SE = 0.000855, p = 0.178 | | | | | |
| BMI-Tg | MR Egger | 223 | 0.302 | 0.065 | 5.45E-06 |
| | Weighted median | 223 | 0.227 | 0.029 | 5.83E-15 |
| | Inverse variance weighted | 223 | 0.203 | 0.022 | 6.79E-20 |
| | Simple mode | 223 | 0.216 | 0.085 | 1.20E-02 |
| | Weighted mode | 223 | 0.239 | 0.049 | 2.45E-06 |
| Test for horizontal pleiotropy: MR Egger intercept = -0.00220, SE = 0.00135, p = 0.105 | | | | | |
| BMI-HDL | MR Egger | 223 | -0.341 | 0.068 | 1.13E-06 |
| | Weighted median | 223 | -0.255 | 0.031 | 1.24E-16 |
| | Inverse variance weighted | 223 | -0.265 | 0.023 | 3.28E-30 |
| | Simple mode | 223 | -0.050 | 0.074 | 5.01E-01 |
| | Weighted mode | 223 | -0.285 | 0.045 | 1.73E-09 |
| Test for horizontal pleiotropy: MR Egger intercept = -0.00169, SE = 0.00141, p = 0.234 | | | | | |
| BMI-WBC | MR Egger | 298 | -0.035 | 0.045 | 4.34E-01 |
| | Weighted median | 298 | 0.031 | 0.011 | 6.15E-03 |
| | Inverse variance weighted | 298 | 0.019 | 0.015 | 2.23E-01 |
| | Simple mode | 298 | 0.028 | 0.037 | 4.46E-01 |
| | Weighted mode | 298 | 0.054 | 0.030 | 7.28E-02 |

| | | | | | |
|---|---|---|---|---|---|
| Test for horizontal pleiotropy: MR Egger intercept = -0.00118, SE = 0.000921, p = 0.202 | | | | | |
| BMI-cortisol | MR Egger | 246 | -0.108 | 0.149 | 4.69E-01 |
| | Weighted median | 246 | -0.068 | 0.075 | 3.63E-01 |
| | Inverse variance weighted | 246 | -0.014 | 0.049 | 7.80E-01 |
| | Simple mode | 246 | -0.115 | 0.207 | 5.78E-01 |
| | Weighted mode | 246 | -0.173 | 0.130 | 1.86E-01 |
| Test for horizontal pleiotropy: MR Egger intercept = -0.00202, SE = 0.00301, p = 0.503 | | | | | |
| BMI-SHBG | MR Egger | 492 | -6.193 | 1.182 | 2.40E-07 |
| | Weighted median | 492 | -6.065 | 0.383 | 1.67E-56 |
| | Inverse variance weighted | 492 | -7.084 | 0.445 | 5.67E-57 |
| | Simple mode | 492 | -6.315 | 1.277 | 1.06E-06 |
| | Weighted mode | 492 | -5.343 | 0.777 | 1.88E-11 |
| Test for horizontal pleiotropy: MR Egger intercept = -0.0156, SE = 0.0191, p = 0.416 | | | | | |
| BMI-folate | MR Egger | 492 | -0.009 | 0.052 | 8.69E-01 |
| | Weighted median | 492 | -0.023 | 0.029 | 4.17E-01 |
| | Inverse variance weighted | 492 | -0.059 | 0.019 | 2.27E-03 |
| | Simple mode | 492 | -0.011 | 0.091 | 9.03E-01 |
| | Weighted mode | 492 | 0.025 | 0.052 | 6.36E-01 |
| Test for horizontal pleiotropy: MR Egger intercept = -0.000891, SE = 0.000836, p = 0.288 | | | | | |
| BMI-RDW | MR Egger | 300 | 0.245 | 0.055 | 1.18E-05 |
| | Weighted median | 300 | 0.169 | 0.021 | 1.67E-15 |
| | Inverse variance weighted | 300 | 0.162 | 0.019 | 1.08E-17 |
| | Simple mode | 300 | 0.192 | 0.060 | 1.53E-03 |
| | Weighted mode | 300 | 0.211 | 0.039 | 1.03E-07 |
| Test for horizontal pleiotropy: MR Egger intercept = -0.00184, SE = 0.001134, p = 0.107 | | | | | |
| BMI-Glu | MR Egger | 64 | 0.107 | 0.035 | 3.05E-03 |
| | Weighted median | 64 | 0.088 | 0.019 | 5.11E-06 |
| | Inverse variance weighted | 64 | 0.093 | 0.015 | 3.08E-10 |
| | Simple mode | 64 | 0.094 | 0.035 | 9.83E-03 |
| | Weighted mode | 64 | 0.089 | 0.022 | 1.87E-04 |
| Test for horizontal pleiotropy: MR Egger intercept = -0.0003952, SE = 0.00084453, p = 0.6415 | | | | | |
| BMI-Fer | MR Egger | 206 | 0.008 | 0.112 | 9.41E-01 |

| | | | | | |
|---|---|---|---|---|---|
| | Weighted median | 206 | 0.108 | 0.057 | 5.91E-02 |
| | Inverse variance weighted | 206 | 0.132 | 0.039 | 6.44E-04 |
| | Simple mode | 206 | 0.196 | 0.180 | 2.77E-01 |
| | Weighted mode | 206 | 0.012 | 0.114 | 9.17E-01 |
| Test for horizontal pleiotropy: MR Egger intercept = 0.002695, SE = 0.002299, p = 0.242 | | | | | |
| BMI-Tes | MR Egger | 205 | -0.262 | 0.112 | 2.05E-02 |
| | Weighted median | 205 | -0.337 | 0.060 | 1.82E-08 |
| | Inverse variance weighted | 205 | -0.147 | 0.041 | 3.61E-04 |
| | Simple mode | 205 | -0.559 | 0.153 | 3.25E-04 |
| | Weighted mode | 205 | -0.440 | 0.091 | 2.71E-06 |
| Test for horizontal pleiotropy: MR Egger intercept = 0.002656, SE = 0.002407, p = 0.271 | | | | | |
| BMI-Baso | MR Egger | 304 | -0.006 | 0.038 | 8.81E-01 |
| | Weighted median | 304 | -0.036 | 0.018 | 4.70E-02 |
| | Inverse variance weighted | 304 | -0.009 | 0.013 | 4.90E-01 |
| | Simple mode | 304 | -0.069 | 0.055 | 2.05E-01 |
| | Weighted mode | 304 | -0.059 | 0.042 | 1.63E-01 |
| Test for horizontal pleiotropy: MR Egger intercept = -7.13E-05, SE = 0.000785, p = 0.928 | | | | | |
| BMI-Mono | MR Egger | 298 | -0.097 | 0.045 | 3.07E-02 |
| | Weighted median | 298 | -0.015 | 0.011 | 1.82E-01 |
| | Inverse variance weighted | 298 | -0.018 | 0.015 | 2.37E-01 |
| | Simple mode | 298 | -0.004 | 0.035 | 9.18E-01 |
| | Weighted mode | 298 | -0.016 | 0.023 | 4.84E-01 |
| Test for horizontal pleiotropy: MR Egger intercept = 0.001740, SE = 0.000926404, p = 0.0613 | | | | | |
| BMI-MCV | MR Egger | 300 | -0.108 | 0.147 | 4.62E-01 |
| | Weighted median | 300 | -0.141 | 0.079 | 7.48E-02 |
| | Inverse variance weighted | 300 | -0.087 | 0.049 | 7.42E-02 |
| | Simple mode | 300 | -0.097 | 0.229 | 6.71E-01 |
| | Weighted mode | 300 | -0.147 | 0.158 | 3.53E-01 |
| Test for horizontal pleiotropy: MR Egger intercept = 0.000459, SE = 0.003011, p = 0.879 | | | | | |
| BMI-Lymph | MR Egger | 298 | -0.076 | 0.048 | 1.11E-01 |
| | Weighted median | 298 | -0.026 | 0.012 | 3.15E-02 |
| | Inverse variance weighted | 298 | 0.020 | 0.016 | 2.20E-01 |
| | Simple mode | 298 | -0.028 | 0.038 | 4.64E-01 |
| | Weighted mode | 298 | -0.048 | 0.022 | 3.20E-02 |
| Test for horizontal pleiotropy: MR Egger intercept =0.002111, SE = 0.0009811, p = 0.032161341 | | | | | |

**Table 4S. 2S-MR results with BMI with biomarkers as exposures and BMI as outcome to assess reverse causality**

| id.exposure | id.outcome | outcome | exposure | method | nsnp | b | se | pval |
|---|---|---|---|---|---|---|---|---|
| **ebi-a-GCST004631** | ukb-a-248 | Body mass index (BMI) \|\| id:ukb-a-248 | Basophil percentage of white cells \|\| id:ebi-a-GCST004631 | MR Egger | 55 | 0.02850274 | 0.03058163 | 0.35555179 |
| **ebi-a-GCST004631** | ukb-a-248 | Body mass index (BMI) \|\| id:ukb-a-248 | Basophil percentage of white cells \|\| id:ebi-a-GCST004631 | Weighted median | 55 | 0.01115946 | 0.01802141 | 0.53576264 |
| **ebi-a-GCST004631** | ukb-a-248 | Body mass index (BMI) \|\| id:ukb-a-248 | Basophil percentage of white cells \|\| id:ebi-a-GCST004631 | Inverse variance weighted | 55 | -0.0133255 | 0.01548404 | 0.38946137 |
| **ebi-a-GCST004631** | ukb-a-248 | Body mass index (BMI) \|\| id:ukb-a-248 | Basophil percentage of white cells \|\| id:ebi-a-GCST004631 | Simple mode | 55 | 0.00553647 | 0.03733532 | 0.88266604 |
| **ebi-a-GCST004631** | ukb-a-248 | Body mass index (BMI) \|\| id:ukb-a-248 | Basophil percentage of white cells \|\| id:ebi-a-GCST004631 | Weighted mode | 55 | -0.0005482 | 0.02054445 | 0.97881084 |
| **id.exposure** | id.outcome | outcome | exposure | method | nsnp | b | se | pval |
| **ieu-a-1012** | ukb-a-248 | Body mass index (BMI) \|\| id:ukb-a-248 | Plasma cortisol \|\| id:ieu-a-1012 | Wald ratio | 1 | -0.0162841 | 0.0285215 | 0.56803913 |
| **id.exposure** | id.outcome | outcome | exposure | method | nsnp | b | se | pval |
| **ieu-a-1050** | ukb-a-248 | Body mass index (BMI) \|\| id:ukb-a-248 | Ferritin \|\| id:ieu-a-1050 | MR Egger | 4 | -0.0692185 | 0.02954547 | 0.14388806 |
| **ieu-a-1050** | ukb-a-248 | Body mass index (BMI) \|\| id:ukb-a-248 | Ferritin \|\| id:ieu-a-1050 | Weighted median | 4 | -0.0458501 | 0.01819113 | 0.01171994 |
| **ieu-a-1050** | ukb-a-248 | Body mass index (BMI) \|\| id:ukb-a-248 | Ferritin \|\| id:ieu-a-1050 | Inverse variance weighted | 4 | -0.0401901 | 0.01571805 | 0.01055975 |
| **ieu-a-1050** | ukb-a-248 | Body mass index (BMI) \|\| id:ukb-a-248 | Ferritin \|\| id:ieu-a-1050 | Simple mode | 4 | -0.040832 | 0.02570683 | 0.21040991 |
| **ieu-a-1050** | ukb-a-248 | Body mass index (BMI) \|\| id:ukb-a-248 | Ferritin \|\| id:ieu-a-1050 | Weighted mode | 4 | -0.0508659 | 0.01904314 | 0.07562034 |

<table>
<tr><th>id.exposure</th><th>id.outcome</th><th>outcome</th><th>exposure</th><th>method</th><th>nsnp</th><th>b</th><th>se</th><th>pval</th></tr>
<tr><td>ukb-b-11349</td><td>ieu-b-40</td><td>body mass index || id:ieu-b-40</td><td>Folate || id:ukb-b-11349</td><td>Wald ratio</td><td>1</td><td>0.04546597</td><td>0.05883832</td><td>0.4396838</td></tr>
<tr><td>id.exposure</td><td>id.outcome</td><td>outcome</td><td>exposure</td><td>method</td><td>nsnp</td><td>b</td><td>se</td><td>pval</td></tr>
<tr><td>ieu-b-114</td><td>ukb-a-248</td><td>Body mass index (BMI) || id:ukb-a-248</td><td>Fasting glucose || id:ieu-b-114</td><td>MR Egger</td><td>30</td><td>-0.0453952</td><td>0.11342853</td><td>0.69203895</td></tr>
<tr><td>ieu-b-114</td><td>ukb-a-248</td><td>Body mass index (BMI) || id:ukb-a-248</td><td>Fasting glucose || id:ieu-b-114</td><td>Weighted median</td><td>30</td><td>0.00266784</td><td>0.03188398</td><td>0.93331616</td></tr>
<tr><td>ieu-b-114</td><td>ukb-a-248</td><td>Body mass index (BMI) || id:ukb-a-248</td><td>Fasting glucose || id:ieu-b-114</td><td>Inverse variance weighted</td><td>30</td><td>-0.0341858</td><td>0.05296448</td><td>0.51863721</td></tr>
<tr><td>ieu-b-114</td><td>ukb-a-248</td><td>Body mass index (BMI) || id:ukb-a-248</td><td>Fasting glucose || id:ieu-b-114</td><td>Simple mode</td><td>30</td><td>-0.01322</td><td>0.06706873</td><td>0.84511555</td></tr>
<tr><td>ieu-b-114</td><td>ukb-a-248</td><td>Body mass index (BMI) || id:ukb-a-248</td><td>Fasting glucose || id:ieu-b-114</td><td>Weighted mode</td><td>30</td><td>0.00016512</td><td>0.02915844</td><td>0.99552048</td></tr>
<tr><td>id.exposure</td><td>id.outcome</td><td>outcome</td><td>exposure</td><td>method</td><td>nsnp</td><td>b</td><td>se</td><td>pval</td></tr>
<tr><td>ieu-a-270</td><td>ukb-a-248</td><td>Body mass index (BMI) || id:ukb-a-248</td><td>Haemoglobin concentration || id:ieu-a-270</td><td>MR Egger</td><td>15</td><td>0.00144021</td><td>0.07587258</td><td>0.98514372</td></tr>
<tr><td>ieu-a-270</td><td>ukb-a-248</td><td>Body mass index (BMI) || id:ukb-a-248</td><td>Haemoglobin concentration || id:ieu-a-270</td><td>Weighted median</td><td>15</td><td>0.01307681</td><td>0.02023195</td><td>0.51805628</td></tr>
<tr><td>ieu-a-270</td><td>ukb-a-248</td><td>Body mass index (BMI) || id:ukb-a-248</td><td>Haemoglobin concentration || id:ieu-a-270</td><td>Inverse variance weighted</td><td>15</td><td>-0.0334432</td><td>0.02660806</td><td>0.20879683</td></tr>
<tr><td>ieu-a-270</td><td>ukb-a-248</td><td>Body mass index (BMI) || id:ukb-a-248</td><td>Haemoglobin concentration || id:ieu-a-270</td><td>Simple mode</td><td>15</td><td>-0.1129022</td><td>0.05448673</td><td>0.05720502</td></tr>
<tr><td>ieu-a-270</td><td>ukb-a-248</td><td>Body mass index (BMI) || id:ukb-a-248</td><td>Haemoglobin concentration || id:ieu-a-270</td><td>Weighted mode</td><td>15</td><td>0.01648048</td><td>0.02138074</td><td>0.45363255</td></tr>
</table>

| id.exposure | id.outcome | outcome | exposure | method | nsnp | b | se | pval |
|---|---|---|---|---|---|---|---|---|
| **ieu-b-103** | ukb-a-248 | Body mass index (BMI) \|\| id:ukb-a-248 | HbA1C \|\| id:ieu-b-103 | MR Egger | 11 | 0.01268313 | 0.08396 | 0.88325885 |
| **ieu-b-103** | ukb-a-248 | Body mass index (BMI) \|\| id:ukb-a-248 | HbA1C \|\| id:ieu-b-103 | Weighted median | 11 | -0.0069654 | 0.03248061 | 0.83019839 |
| **ieu-b-103** | ukb-a-248 | Body mass index (BMI) \|\| id:ukb-a-248 | HbA1C \|\| id:ieu-b-103 | Inverse variance weighted | 11 | 0.0283815 | 0.03446064 | 0.41017145 |
| **ieu-b-103** | ukb-a-248 | Body mass index (BMI) \|\| id:ukb-a-248 | HbA1C \|\| id:ieu-b-103 | Simple mode | 11 | -0.0257779 | 0.06293863 | 0.69075483 |
| **ieu-b-103** | ukb-a-248 | Body mass index (BMI) \|\| id:ukb-a-248 | HbA1C \|\| id:ieu-b-103 | Weighted mode | 11 | -0.0208929 | 0.03914067 | 0.60514756 |
| **id.exposure** | id.outcome | outcome | exposure | method | nsnp | b | se | pval |
| **ieu-a-275** | ukb-a-248 | Body mass index (BMI) \|\| id:ukb-a-248 | Red blood cell count \|\| id:ieu-a-275 | MR Egger | 26 | 0.26135491 | 0.14952437 | 0.09326378 |
| **ieu-a-275** | ukb-a-248 | Body mass index (BMI) \|\| id:ukb-a-248 | Red blood cell count \|\| id:ieu-a-275 | Weighted median | 26 | 0.08455861 | 0.04638846 | 0.06832803 |
| **ieu-a-275** | ukb-a-248 | Body mass index (BMI) \|\| id:ukb-a-248 | Red blood cell count \|\| id:ieu-a-275 | Inverse variance weighted | 26 | -0.0072778 | 0.05705404 | 0.89849796 |
| **ieu-a-275** | ukb-a-248 | Body mass index (BMI) \|\| id:ukb-a-248 | Red blood cell count \|\| id:ieu-a-275 | Simple mode | 26 | 0.09054068 | 0.07897524 | 0.26246646 |
| **ieu-a-275** | ukb-a-248 | Body mass index (BMI) \|\| id:ukb-a-248 | Red blood cell count \|\| id:ieu-a-275 | Weighted mode | 26 | 0.09962869 | 0.04750758 | 0.04626112 |

| id.exposure | id.outcome | outcome | exposure | method | nsnp | b | se | pval |
|---|---|---|---|---|---|---|---|---|
| **ebi-a-GCST005068** | ieu-b-40 | body mass index \|\| id:ieu-b-40 | LDL cholesterol \|\| id:ebi-a-GCST005068 | MR Egger | 4 | 0.01267769 | 0.05816618 | 0.84767998 |
| **ebi-a-GCST005068** | ieu-b-40 | body mass index \|\| id:ieu-b-40 | LDL cholesterol \|\| id:ebi-a-GCST005068 | Weighted median | 4 | -0.0332107 | 0.01134854 | 0.00342875 |
| **ebi-a-GCST005068** | ieu-b-40 | body mass index \|\| id:ieu-b-40 | LDL cholesterol \|\| id:ebi-a-GCST005068 | Inverse variance weighted | 4 | -0.0320938 | 0.00863712 | 0.00020257 |
| **ebi-a-GCST005068** | ieu-b-40 | body mass index \|\| id:ieu-b-40 | LDL cholesterol \|\| id:ebi-a-GCST005068 | Simple mode | 4 | -0.036639 | 0.01673254 | 0.11628833 |
| **ebi-a-GCST005068** | ieu-b-40 | body mass index \|\| id:ieu-b-40 | LDL cholesterol \|\| id:ebi-a-GCST005068 | Weighted mode | 4 | -0.034971 | 0.01482963 | 0.09956398 |
| **id.exposure** | id.outcome | outcome | exposure | method | nsnp | b | se | pval |
| **ebi-a-GCST90002336** | ukb-a-248 | Body mass index (BMI) \|\| id:ukb-a-248 | Mean corpuscular volume \|\| id:ebi-a-GCST90002336 | MR Egger | 9 | 0.01627788 | 0.03256415 | 0.63249338 |
| **ebi-a-GCST90002336** | ukb-a-248 | Body mass index (BMI) \|\| id:ukb-a-248 | Mean corpuscular volume \|\| id:ebi-a-GCST90002336 | Weighted median | 9 | -0.0169769 | 0.01096864 | 0.12167813 |
| **ebi-a-GCST90002336** | ukb-a-248 | Body mass index (BMI) \|\| id:ukb-a-248 | Mean corpuscular volume \|\| id:ebi-a-GCST90002336 | Inverse variance weighted | 9 | -0.0212777 | 0.01246612 | 0.08785201 |
| **ebi-a-GCST90002336** | ukb-a-248 | Body mass index (BMI) \|\| id:ukb-a-248 | Mean corpuscular volume \|\| id:ebi-a-GCST90002336 | Simple mode | 9 | -0.0220168 | 0.01637922 | 0.21575632 |
| **ebi-a-GCST90002336** | ukb-a-248 | Body mass index (BMI) \|\| id:ukb-a-248 | Mean corpuscular volume \|\| id:ebi-a-GCST90002336 | Weighted mode | 9 | -0.0231087 | 0.01392193 | 0.13552035 |

<table>
<tr><th>id.exposure</th><th>id.outcome</th><th>outcome</th><th>exposure</th><th>method</th><th>nsnp</th><th>b</th><th>se</th><th>pval</th></tr>
<tr><td>**ieu-a-1008**</td><td>ukb-a-248</td><td>Body mass index (BMI) || id:ukb-a-248</td><td>Platelet count || id:ieu-a-1008</td><td>MR Egger</td><td>32</td><td>-0.0006727</td><td>0.00070143</td><td>0.34517459</td></tr>
<tr><td>**ieu-a-1008**</td><td>ukb-a-248</td><td>Body mass index (BMI) || id:ukb-a-248</td><td>Platelet count || id:ieu-a-1008</td><td>Weighted median</td><td>32</td><td>0.00013472</td><td>0.00021428</td><td>0.52954228</td></tr>
<tr><td>**ieu-a-1008**</td><td>ukb-a-248</td><td>Body mass index (BMI) || id:ukb-a-248</td><td>Platelet count || id:ieu-a-1008</td><td>Inverse variance weighted</td><td>32</td><td>-0.0003268</td><td>0.00024878</td><td>0.18895678</td></tr>
<tr><td>**ieu-a-1008**</td><td>ukb-a-248</td><td>Body mass index (BMI) || id:ukb-a-248</td><td>Platelet count || id:ieu-a-1008</td><td>Simple mode</td><td>32</td><td>9.63E-05</td><td>0.00034761</td><td>0.78352525</td></tr>
<tr><td>**ieu-a-1008**</td><td>ukb-a-248</td><td>Body mass index (BMI) || id:ukb-a-248</td><td>Platelet count || id:ieu-a-1008</td><td>Weighted mode</td><td>32</td><td>0.00011659</td><td>0.00024456</td><td>0.63688891</td></tr>
<tr><td>**id.exposure**</td><td>id.outcome</td><td>outcome</td><td>exposure</td><td>method</td><td>nsnp</td><td>b</td><td>se</td><td>pval</td></tr>
<tr><td>**ieu-a-275**</td><td>ukb-a-248</td><td>Body mass index (BMI) || id:ukb-a-248</td><td>Red blood cell count || id:ieu-a-275</td><td>MR Egger</td><td>26</td><td>0.26135491</td><td>0.14952437</td><td>0.09326378</td></tr>
<tr><td>**ieu-a-275**</td><td>ukb-a-248</td><td>Body mass index (BMI) || id:ukb-a-248</td><td>Red blood cell count || id:ieu-a-275</td><td>Weighted median</td><td>26</td><td>0.08455861</td><td>0.04591029</td><td>0.06550111</td></tr>
<tr><td>**ieu-a-275**</td><td>ukb-a-248</td><td>Body mass index (BMI) || id:ukb-a-248</td><td>Red blood cell count || id:ieu-a-275</td><td>Inverse variance weighted</td><td>26</td><td>-0.0072778</td><td>0.05705404</td><td>0.89849796</td></tr>
<tr><td>**ieu-a-275**</td><td>ukb-a-248</td><td>Body mass index (BMI) || id:ukb-a-248</td><td>Red blood cell count || id:ieu-a-275</td><td>Simple mode</td><td>26</td><td>0.09054068</td><td>0.07653474</td><td>0.24793721</td></tr>
<tr><td>**ieu-a-275**</td><td>ukb-a-248</td><td>Body mass index (BMI) || id:ukb-a-248</td><td>Red blood cell count || id:ieu-a-275</td><td>Weighted mode</td><td>26</td><td>0.09962869</td><td>0.04605998</td><td>0.04030183</td></tr>
</table>

| id.exposure | id.outcome | outcome | exposure | method | nsnp | b | se | pval |
|---|---|---|---|---|---|---|---|---|
| **ebi-a-GCST006804** | ukb-a-248 | Body mass index (BMI) \|\| id:ukb-a-248 | Red cell distribution width \|\| id:ebi-a-GCST006804 | MR Egger | 122 | -0.0341912 | 0.03423066 | 0.31987855 |
| **ebi-a-GCST006804** | ukb-a-248 | Body mass index (BMI) \|\| id:ukb-a-248 | Red cell distribution width \|\| id:ebi-a-GCST006804 | Weighted median | 122 | 0.01563428 | 0.00970492 | 0.10718764 |
| **ebi-a-GCST006804** | ukb-a-248 | Body mass index (BMI) \|\| id:ukb-a-248 | Red cell distribution width \|\| id:ebi-a-GCST006804 | Inverse variance weighted | 122 | 0.0300081 | 0.01723486 | 0.08166104 |
| **ebi-a-GCST006804** | ukb-a-248 | Body mass index (BMI) \|\| id:ukb-a-248 | Red cell distribution width \|\| id:ebi-a-GCST006804 | Simple mode | 122 | 0.01857783 | 0.02026892 | 0.3611926 |
| **ebi-a-GCST006804** | ukb-a-248 | Body mass index (BMI) \|\| id:ukb-a-248 | Red cell distribution width \|\| id:ebi-a-GCST006804 | Weighted mode | 122 | 0.01128028 | 0.01217303 | 0.35594689 |
| **id.exposure** | id.outcome | outcome | exposure | method | nsnp | b | se | pval |
| **ieu-a-302** | ukb-a-248 | Body mass index (BMI) \|\| id:ukb-a-248 | Triglycerides \|\| id:ieu-a-302 | MR Egger | 55 | -0.0445751 | 0.03601334 | 0.22126815 |
| **ieu-a-302** | ukb-a-248 | Body mass index (BMI) \|\| id:ukb-a-248 | Triglycerides \|\| id:ieu-a-302 | Weighted median | 55 | -0.0315603 | 0.01590831 | 0.04726817 |
| **ieu-a-302** | ukb-a-248 | Body mass index (BMI) \|\| id:ukb-a-248 | Triglycerides \|\| id:ieu-a-302 | Inverse variance weighted | 55 | -0.0214287 | 0.02233545 | 0.33735629 |
| **ieu-a-302** | ukb-a-248 | Body mass index (BMI) \|\| id:ukb-a-248 | Triglycerides \|\| id:ieu-a-302 | Simple mode | 55 | -0.0456892 | 0.02952211 | 0.1275531 |
| **ieu-a-302** | ukb-a-248 | Body mass index (BMI) \|\| id:ukb-a-248 | Triglycerides \|\| id:ieu-a-302 | Weighted mode | 55 | -0.0297723 | 0.01209893 | 0.01709375 |

| id.exposure | id.outcome | outcome | exposure | method | nsnp | b | se | pval |
|---|---|---|---|---|---|---|---|---|
| **ieu-b-30** | ukb-a-248 | Body mass index (BMI) \|\| id:ukb-a-248 | white blood cell count \|\| id:ieu-b-30 | MR Egger | 475 | -0.0347487 | 0.02445633 | 0.15602056 |
| **ieu-b-30** | ukb-a-248 | Body mass index (BMI) \|\| id:ukb-a-248 | white blood cell count \|\| id:ieu-b-30 | Weighted median | 475 | -0.0307482 | 0.01204555 | 0.01069025 |
| **ieu-b-30** | ukb-a-248 | Body mass index (BMI) \|\| id:ukb-a-248 | white blood cell count \|\| id:ieu-b-30 | Inverse variance weighted | 475 | -0.040535 | 0.01168163 | 0.0005205 |
| **ieu-b-30** | ukb-a-248 | Body mass index (BMI) \|\| id:ukb-a-248 | white blood cell count \|\| id:ieu-b-30 | Simple mode | 475 | 0.00133145 | 0.03782675 | 0.97193623 |
| **ieu-b-30** | ukb-a-248 | Body mass index (BMI) \|\| id:ukb-a-248 | white blood cell count \|\| id:ieu-b-30 | Weighted mode | 475 | -0.0184754 | 0.02085621 | 0.37614876 |

| id.exposure | id.outcome | outcome | exposure | method | nsnp | b | se | pval |
|---|---|---|---|---|---|---|---|---|
| **ukb-d-30830_raw** | ieu-b-40 | body mass index \|\| id:ieu-b-40 | SHBG \|\| id:ukb-d-30830_raw | MR Egger | 135 | 0.00121356 | 0.00180979 | 0.50366715 |
| **ukb-d-30830_raw** | ieu-b-40 | body mass index \|\| id:ieu-b-40 | SHBG \|\| id:ukb-d-30830_raw | Weighted median | 135 | 0.00102542 | 0.00037386 | 0.00609279 |
| **ukb-d-30830_raw** | ieu-b-40 | body mass index \|\| id:ieu-b-40 | SHBG \|\| id:ukb-d-30830_raw | Inverse variance weighted | 135 | -0.002129 | 0.001153 | 0.06482013 |
| **ukb-d-30830_raw** | ieu-b-40 | body mass index \|\| id:ieu-b-40 | SHBG \|\| id:ukb-d-30830_raw | Simple mode | 135 | -0.0011256 | 0.00124723 | 0.36841147 |

#### Table 5S. 2S-MR results with vigorous physical activity as exposure and blood biomarkers as outcomes

| id.exposure | id.outcome | outcome | exposure | method | nsnp | b | se | pval |
|---|---|---|---|---|---|---|---|---|
| **ebi-a-GCST006098** | ebi-a-GCST005068 | LDL cholesterol \|\| id:ebi-a-GCST005068 | Vigorous physical activity \|\| id:ebi-a-GCST006098 | MR Egger | 7 | 4.55735741 | 5.39758261 | 0.43700589 |
| **ebi-a-GCST006098** | ebi-a-GCST005068 | LDL cholesterol \|\| id:ebi-a-GCST005068 | Vigorous physical activity \|\| id:ebi-a-GCST006098 | Weighted median | 7 | -0.6208293 | 0.83843447 | 0.45901939 |
| **ebi-a-GCST006098** | ebi-a-GCST005068 | LDL cholesterol \|\| id:ebi-a-GCST005068 | Vigorous physical activity \|\| id:ebi-a-GCST006098 | Inverse variance weighted | 7 | -0.2426177 | 0.66016146 | 0.71323661 |
| **ebi-a-GCST006098** | ebi-a-GCST005068 | LDL cholesterol \|\| id:ebi-a-GCST005068 | Vigorous physical activity \|\| id:ebi-a-GCST006098 | Simple mode | 7 | -1.231375 | 1.53251313 | 0.45233386 |
| **ebi-a-GCST006098** | ebi-a-GCST005068 | LDL cholesterol \|\| id:ebi-a-GCST005068 | Vigorous physical activity \|\| id:ebi-a-GCST006098 | Weighted mode | 7 | -1.0821029 | 1.01739534 | 0.32843007 |
| **ebi-a-GCST006098** | ieu-a-1050 | Ferritin \|\| id:ieu-a-1050 | Vigorous physical activity \|\| id:ebi-a-GCST006098 | MR Egger | 5 | -5.2248204 | 5.49515103 | 0.41184823 |
| **ebi-a-GCST006098** | ieu-a-1050 | Ferritin \|\| id:ieu-a-1050 | Vigorous physical activity \|\| id:ebi-a-GCST006098 | Weighted median | 5 | -0.7040971 | 0.64752149 | 0.27687222 |
| **ebi-a-GCST006098** | ieu-a-1050 | Ferritin \|\| id:ieu-a-1050 | Vigorous physical activity \|\| id:ebi-a-GCST006098 | Inverse variance weighted | 5 | -0.1408587 | 0.76723061 | 0.85433225 |
| **ebi-a-GCST006098** | ieu-a-1050 | Ferritin \|\| id:ieu-a-1050 | Vigorous physical activity \|\| id:ebi-a-GCST006098 | Simple mode | 5 | -0.8515134 | 0.84328604 | 0.36973273 |
| **ebi-a-GCST006098** | ieu-a-1050 | Ferritin \|\| id:ieu-a-1050 | Vigorous physical activity \|\| id:ebi-a-GCST006098 | Weighted mode | 5 | -0.9327437 | 0.76915852 | 0.29197296 |
| **ebi-a-GCST006098** | ieu-a-302 | Triglycerides \|\| id:ieu-a-302 | Vigorous physical activity \|\| id:ebi-a-GCST006098 | MR Egger | 5 | -1.4775783 | 1.33435404 | 0.34895532 |
| **ebi-a-GCST006098** | ieu-a-302 | Triglycerides \|\| id:ieu-a-302 | Vigorous physical activity \|\| id:ebi-a-GCST006098 | Weighted median | 5 | -0.1485007 | 0.26200258 | 0.5708563 |
| **ebi-a-GCST006098** | ieu-a-302 | Triglycerides \|\| id:ieu-a-302 | Vigorous physical activity \|\| id:ebi-a-GCST006098 | Inverse variance weighted | 5 | -0.0745059 | 0.22114262 | 0.73618224 |
| **ebi-a-GCST006098** | ieu-a-302 | Triglycerides \|\| id:ieu-a-302 | Vigorous physical activity \|\| id:ebi-a-GCST006098 | Simple mode | 5 | -0.1338663 | 0.36851473 | 0.73479489 |
| **ebi-a-GCST006098** | ieu-a-302 | Triglycerides \|\| id:ieu-a-302 | Vigorous physical activity \|\| id:ebi-a-GCST006098 | Weighted mode | 5 | -0.1563444 | 0.32885888 | 0.65928919 |
| **ebi-a-GCST006098** | ukb-b-11349 | Folate \|\| id:ukb-b-11349 | Vigorous physical activity \|\| id:ebi-a-GCST006098 | MR Egger | 7 | 0.44992252 | 2.05817378 | 0.83560206 |
| **ebi-a-GCST006098** | ukb-b-11349 | Folate \|\| id:ukb-b-11349 | Vigorous physical activity \|\| id:ebi-a-GCST006098 | Weighted median | 7 | -0.2016639 | 0.32005364 | 0.52863309 |
| **ebi-a-GCST006098** | ukb-b-11349 | Folate \|\| id:ukb-b-11349 | Vigorous physical activity \|\| id:ebi-a-GCST006098 | Inverse variance weighted | 7 | -0.2577298 | 0.24461528 | 0.29206017 |
| **ebi-a-GCST006098** | ukb-b-11349 | Folate \|\| id:ukb-b-11349 | Vigorous physical activity \|\| id:ebi-a-GCST006098 | Simple mode | 7 | 0.0568742 | 0.46702435 | 0.90704956 |
| **ebi-a-GCST006098** | ukb-b-11349 | Folate \|\| id:ukb-b-11349 | Vigorous physical activity \|\| id:ebi-a-GCST006098 | Weighted mode | 7 | 0.07601041 | 0.46533968 | 0.87561065 |

| | | | | | | | | |
|---|---|---|---|---|---|---|---|---|
| **ebi-a-GCST006098** | **ukb-d-30070_irnt** | **Red blood cell (erythrocyte) distribution width \|\| id:ukb-d-30070_irnt** | **Vigorous physical activity \|\| id:ebi-a-GCST006098** | **MR Egger** | **7** | **1.57820562** | **1.79570092** | **0.41969132** |
| **ebi-a-GCST006098** | ukb-d-30070_irnt | Red blood cell (erythrocyte) distribution width \|\| id:ukb-d-30070_irnt | Vigorous physical activity \|\| id:ebi-a-GCST006098 | Weighted median | 7 | -0.0549663 | 0.14390626 | 0.70249175 |
| **ebi-a-GCST006098** | ukb-d-30070_irnt | Red blood cell (erythrocyte) distribution width \|\| id:ukb-d-30070_irnt | Vigorous physical activity \|\| id:ebi-a-GCST006098 | Inverse variance weighted | 7 | -0.2321729 | 0.21329882 | 0.27638026 |
| **ebi-a-GCST006098** | ukb-d-30070_irnt | Red blood cell (erythrocyte) distribution width \|\| id:ukb-d-30070_irnt | Vigorous physical activity \|\| id:ebi-a-GCST006098 | Simple mode | 7 | -0.0408618 | 0.17733468 | 0.82541927 |
| **ebi-a-GCST006098** | ukb-d-30070_irnt | Red blood cell (erythrocyte) distribution width \|\| id:ukb-d-30070_irnt | Vigorous physical activity \|\| id:ebi-a-GCST006098 | Weighted mode | 7 | -0.0920344 | 0.18214756 | 0.63138589 |
| **ebi-a-GCST006098** | ukb-d-30830_irnt | SHBG \|\| id:ukb-d-30830_irnt | Vigorous physical activity \|\| id:ebi-a-GCST006098 | MR Egger | 7 | 0.46503747 | 1.9737129 | 0.82307752 |
| **ebi-a-GCST006098** | ukb-d-30830_irnt | SHBG \|\| id:ukb-d-30830_irnt | Vigorous physical activity \|\| id:ebi-a-GCST006098 | Weighted median | 7 | 0.11870029 | 0.15191489 | 0.43459052 |
| **ebi-a-GCST006098** | ukb-d-30830_irnt | SHBG \|\| id:ukb-d-30830_irnt | Vigorous physical activity \|\| id:ebi-a-GCST006098 | Inverse variance weighted | 7 | 0.33282495 | 0.21369539 | 0.11935806 |
| **ebi-a-GCST006098** | ukb-d-30830_irnt | SHBG \|\| id:ukb-d-30830_irnt | Vigorous physical activity \|\| id:ebi-a-GCST006098 | Simple mode | 7 | 0.07719013 | 0.19009622 | 0.6987848 |
| **ebi-a-GCST006098** | ukb-d-30830_irnt | SHBG \|\| id:ukb-d-30830_irnt | Vigorous physical activity \|\| id:ebi-a-GCST006098 | Weighted mode | 7 | 0.08827653 | 0.18228932 | 0.64536599 |

#### Tests for horizontal pleiotropy

| id.exposure | id.outcome | outcome | exposure | Egger intercept | se | pval |
|---|---|---|---|---|---|---|
| **ebi-a-GCST006098** | ebi-a-GCST005068 | LDL cholesterol \|\| id:ebi-a-GCST005068 | Vigorous physical activity \|\| id:ebi-a-GCST006098 | -0.0463331 | 0.05171055 | 0.41130582 |
| **ebi-a-GCST006098** | ieu-a-1050 | Ferritin \|\| id:ieu-a-1050 | Vigorous physical activity \|\| id:ebi-a-GCST006098 | 0.04824757 | 0.05162226 | 0.41892633 |
| **ebi-a-GCST006098** | ieu-a-302 | Triglycerides \|\| id:ieu-a-302 | Vigorous physical activity \|\| id:ebi-a-GCST006098 | 0.01391277 | 0.01304839 | 0.36450721 |
| **ebi-a-GCST006098** | ukb-b-11349 | Folate \|\| id:ukb-b-11349 | Vigorous physical activity \|\| id:ebi-a-GCST006098 | -0.0065813 | 0.01900584 | 0.74322824 |
| **ebi-a-GCST006098** | ukb-d-30070_irnt | Red blood cell (erythrocyte) distribution width \|\| id:ukb-d-30070_irnt | Vigorous physical activity \|\| id:ebi-a-GCST006098 | -0.0168346 | 0.01658053 | 0.35653574 |
| **ebi-a-GCST006098** | ukb-d-30830_irnt | SHBG \|\| id:ukb-d-30830_irnt | Vigorous physical activity \|\| id:ebi-a-GCST006098 | -0.0012295 | 0.01822548 | 0.94882818 |

### Table 6S. 2S-MR results with vigorous physical activity as exposure and blood biomarkers as outcomes

| id.exposure | id.outcome | outcome | exposure | method | nsnp | b | se | pval |
|---|---|---|---|---|---|---|---|---|
| **ebi-a-GCST006098** | ukb-b-10217 | Sweets intake \|\| id:ukb-b-10217 | Vigorous physical activity \|\| id:ebi-a-GCST006098 | MR Egger | 7 | -0.0903539 | 1.5078935 | 0.95454011 |
| **ebi-a-GCST006098** | ukb-b-10217 | Sweets intake \|\| id:ukb-b-10217 | Vigorous physical activity \|\| id:ebi-a-GCST006098 | Weighted median | 7 | -0.2367422 | 0.21224606 | 0.26467304 |
| **ebi-a-GCST006098** | ukb-b-10217 | Sweets intake \|\| id:ukb-b-10217 | Vigorous physical activity \|\| id:ebi-a-GCST006098 | Inverse variance weighted | 7 | -0.1760505 | 0.17921106 | 0.32592043 |
| **ebi-a-GCST006098** | ukb-b-10217 | Sweets intake \|\| id:ukb-b-10217 | Vigorous physical activity \|\| id:ebi-a-GCST006098 | Simple mode | 7 | -0.2770321 | 0.31802813 | 0.41719033 |
| **ebi-a-GCST006098** | ukb-b-10217 | Sweets intake \|\| id:ukb-b-10217 | Vigorous physical activity \|\| id:ebi-a-GCST006098 | Weighted mode | 7 | -0.2596458 | 0.31161898 | 0.43662814 |
| **ebi-a-GCST006098** | ukb-b-11679 | Type of special diet followed: Vegetarian \|\| id:ukb-b-11679 | Vigorous physical activity \|\| id:ebi-a-GCST006098 | MR Egger | 6 | 0.30989332 | 0.88272515 | 0.74325054 |
| **ebi-a-GCST006098** | ukb-b-11679 | Type of special diet followed: Vegetarian \|\| id:ukb-b-11679 | Vigorous physical activity \|\| id:ebi-a-GCST006098 | Weighted median | 6 | 0.02203046 | 0.0642637 | 0.73173884 |
| **ebi-a-GCST006098** | ukb-b-11679 | Type of special diet followed: Vegetarian \|\| id:ukb-b-11679 | Vigorous physical activity \|\| id:ebi-a-GCST006098 | Inverse variance weighted | 6 | 0.02308089 | 0.04874316 | 0.63584178 |
| **ebi-a-GCST006098** | ukb-b-11679 | Type of special diet followed: Vegetarian \|\| id:ukb-b-11679 | Vigorous physical activity \|\| id:ebi-a-GCST006098 | Simple mode | 6 | 0.02147452 | 0.10369298 | 0.84410404 |
| **ebi-a-GCST006098** | ukb-b-11679 | Type of special diet followed: Vegetarian \|\| id:ukb-b-11679 | Vigorous physical activity \|\| id:ebi-a-GCST006098 | Weighted mode | 6 | 0.02451679 | 0.09506696 | 0.80677012 |
| **ebi-a-GCST006098** | ukb-b-1996 | Salad / raw vegetable intake \|\| id:ukb-b-1996 | Vigorous physical activity \|\| id:ebi-a-GCST006098 | MR Egger | 7 | 0.66619702 | 1.2339482 | 0.61244007 |
| **ebi-a-GCST006098** | ukb-b-1996 | Salad / raw vegetable intake \|\| id:ukb-b-1996 | Vigorous physical activity \|\| id:ebi-a-GCST006098 | Weighted median | 7 | 0.5139604 | 0.10299379 | 6.03E-07 |
| **ebi-a-GCST006098** | ukb-b-1996 | Salad / raw vegetable intake \|\| id:ukb-b-1996 | Vigorous physical activity \|\| id:ebi-a-GCST006098 | Inverse variance weighted | 7 | 0.50448517 | 0.13388668 | 0.00016456 |
| **ebi-a-GCST006098** | ukb-b-1996 | Salad / raw vegetable intake \|\| id:ukb-b-1996 | Vigorous physical activity \|\| id:ebi-a-GCST006098 | Simple mode | 7 | 0.62702954 | 0.14130406 | 0.00438784 |

| | | | | | | | | |
|---|---|---|---|---|---|---|---|---|
| **ebi-a-GCST006098** | **ukb-b-1996** | **Salad / raw vegetable intake \|\| id:ukb-b-1996** | **Vigorous physical activity \|\| id:ebi-a-GCST006098** | **Weighted mode** | **7** | **0.58940834** | **0.1467562** | **0.00698862** |
| **ebi-a-GCST006098** | ukb-b-2209 | Oily fish intake \|\| id:ukb-b-2209 | Vigorous physical activity \|\| id:ebi-a-GCST006098 | MR Egger | 7 | 0.95472369 | 2.00996591 | 0.65481403 |
| **ebi-a-GCST006098** | ukb-b-2209 | Oily fish intake \|\| id:ukb-b-2209 | Vigorous physical activity \|\| id:ebi-a-GCST006098 | Weighted median | 7 | 0.53975424 | 0.15310124 | 0.00042273 |
| **ebi-a-GCST006098** | ukb-b-2209 | Oily fish intake \|\| id:ukb-b-2209 | Vigorous physical activity \|\| id:ebi-a-GCST006098 | Inverse variance weighted | 7 | 0.48170818 | 0.21898245 | 0.02782414 |
| **ebi-a-GCST006098** | ukb-b-2209 | Oily fish intake \|\| id:ukb-b-2209 | Vigorous physical activity \|\| id:ebi-a-GCST006098 | Simple mode | 7 | 0.82403974 | 0.20581323 | 0.00708805 |
| **ebi-a-GCST006098** | ukb-b-2209 | Oily fish intake \|\| id:ukb-b-2209 | Vigorous physical activity \|\| id:ebi-a-GCST006098 | Weighted mode | 7 | 0.78663663 | 0.29210032 | 0.03590724 |
| **ebi-a-GCST006098** | ukb-b-3881 | Fresh fruit intake \|\| id:ukb-b-3881 | Vigorous physical activity \|\| id:ebi-a-GCST006098 | MR Egger | 7 | 1.30879467 | 1.11224869 | 0.29227451 |
| **ebi-a-GCST006098** | ukb-b-3881 | Fresh fruit intake \|\| id:ukb-b-3881 | Vigorous physical activity \|\| id:ebi-a-GCST006098 | Weighted median | 7 | 0.46001672 | 0.08901784 | 2.37E-07 |
| **ebi-a-GCST006098** | ukb-b-3881 | Fresh fruit intake \|\| id:ukb-b-3881 | Vigorous physical activity \|\| id:ebi-a-GCST006098 | Inverse variance weighted | 7 | 0.38633487 | 0.12863774 | 0.00267088 |
| **ebi-a-GCST006098** | ukb-b-3881 | Fresh fruit intake \|\| id:ukb-b-3881 | Vigorous physical activity \|\| id:ebi-a-GCST006098 | Simple mode | 7 | 0.59733916 | 0.1169325 | 0.00220313 |
| **ebi-a-GCST006098** | ukb-b-3881 | Fresh fruit intake \|\| id:ukb-b-3881 | Vigorous physical activity \|\| id:ebi-a-GCST006098 | Weighted mode | 7 | 0.58037912 | 0.12022717 | 0.00291812 |
| **ebi-a-GCST006098** | ukb-b-4616 | Nap during day \|\| id:ukb-b-4616 | Vigorous physical activity \|\| id:ebi-a-GCST006098 | MR Egger | 7 | -1.8168346 | 0.8575858 | 0.08766702 |

| | | | | | | | | |
|---|---|---|---|---|---|---|---|---|
| **ebi-a-GCST006098** | **ukb-b-4616** | **Nap during day \|\| id:ukb-b-4616** | **Vigorous physical activity \|\| id:ebi-a-GCST006098** | **Weighted median** | **7** | **-0.3203349** | **0.09563435** | **0.00080934** |
| **ebi-a-GCST006098** | ukb-b-4616 | Nap during day \|\| id:ukb-b-4616 | Vigorous physical activity \|\| id:ebi-a-GCST006098 | Inverse variance weighted | 7 | -0.1836272 | 0.12240112 | 0.1335603 |
| **ebi-a-GCST006098** | ukb-b-4616 | Nap during day \|\| id:ukb-b-4616 | Vigorous physical activity \|\| id:ebi-a-GCST006098 | Simple mode | 7 | -0.3872459 | 0.1272748 | 0.02272693 |
| **ebi-a-GCST006098** | ukb-b-4616 | Nap during day \|\| id:ukb-b-4616 | Vigorous physical activity \|\| id:ebi-a-GCST006098 | Weighted mode | 7 | -0.3872459 | 0.11249117 | 0.01375948 |
| **ebi-a-GCST006098** | ukb-b-6324 | Processed meat intake \|\| id:ukb-b-6324 | Vigorous physical activity \|\| id:ebi-a-GCST006098 | MR Egger | 7 | 0.06859678 | 1.00946468 | 0.94845629 |
| **ebi-a-GCST006098** | ukb-b-6324 | Processed meat intake \|\| id:ukb-b-6324 | Vigorous physical activity \|\| id:ebi-a-GCST006098 | Weighted median | 7 | -0.6292381 | 0.13853861 | 5.57E-06 |
| **ebi-a-GCST006098** | ukb-b-6324 | Processed meat intake \|\| id:ukb-b-6324 | Vigorous physical activity \|\| id:ebi-a-GCST006098 | Inverse variance weighted | 7 | -0.5496216 | 0.11346205 | 1.27E-06 |
| **ebi-a-GCST006098** | ukb-b-6324 | Processed meat intake \|\| id:ukb-b-6324 | Vigorous physical activity \|\| id:ebi-a-GCST006098 | Simple mode | 7 | -0.6879646 | 0.21816478 | 0.01972901 |
| **ebi-a-GCST006098** | ukb-b-6324 | Processed meat intake \|\| id:ukb-b-6324 | Vigorous physical activity \|\| id:ebi-a-GCST006098 | Weighted mode | 7 | -0.6939066 | 0.21655974 | 0.01850116 |

**Tests for horizontal pleiotropy**

| id.exposure | id.outcome | outcome | exposure | Egger intercept | se | pval |
|---|---|---|---|---|---|---|
| **ebi-a-GCST006098** | ukb-b-10217 | Sweets intake \|\| id:ukb-b-10217 | Vigorous physical activity \|\| id:ebi-a-GCST006098 | -0.000797 | 0.01392445 | 0.95657282 |
| **ebi-a-GCST006098** | ukb-b-11679 | Type of special diet followed: Vegetarian \|\| id:ukb-b-11679 | Vigorous physical activity \|\| id:ebi-a-GCST006098 | -0.0025594 | 0.00786299 | 0.76112246 |
| **ebi-a-GCST006098** | ukb-b-1996 | Salad / raw vegetable intake \|\| id:ukb-b-1996 | Vigorous physical activity \|\| id:ebi-a-GCST006098 | -0.0015039 | 0.01139436 | 0.90014308 |
| **ebi-a-GCST006098** | ukb-b-2209 | Oily fish intake \|\| id:ukb-b-2209 | Vigorous physical activity \|\| id:ebi-a-GCST006098 | -0.0043991 | 0.01856093 | 0.82205339 |
| **ebi-a-GCST006098** | ukb-b-3881 | Fresh fruit intake \|\| id:ukb-b-3881 | Vigorous physical activity \|\| id:ebi-a-GCST006098 | -0.0085789 | 0.01027085 | 0.44163822 |
| **ebi-a-GCST006098** | ukb-b-4616 | Nap during day \|\| id:ukb-b-4616 | Vigorous physical activity \|\| id:ebi-a-GCST006098 | 0.01518897 | 0.00791926 | 0.11322178 |
| **ebi-a-GCST006098** | ukb-b-6324 | Processed meat intake \|\| id:ukb-b-6324 | Vigorous physical activity \|\| id:ebi-a-GCST006098 | -0.0057496 | 0.009322 | 0.56437631 |

**Table 7S**. **2S-MR with healthy/unhealthy dietary habits as exposures and vigorous physical activity as outcome to assess reverse causality**

| id.exposure | id.outcome | outcome | exposure | method | nsnp | b | se | pval |
|---|---|---|---|---|---|---|---|---|
| **ukb-b-1996** | ebi-a-GCST006098 | Vigorous physical activity \|\| id:ebi-a-GCST006098 | Salad / raw vegetable intake \|\| id:ukb-b-1996 | MR Egger | 20 | 0.3848011 | 0.33578299 | 0.26681018 |
| **ukb-b-1996** | ebi-a-GCST006098 | Vigorous physical activity \|\| id:ebi-a-GCST006098 | Salad / raw vegetable intake \|\| id:ukb-b-1996 | Weighted median | 20 | 0.27257002 | 0.06073408 | 7.19E-06 |
| **ukb-b-1996** | ebi-a-GCST006098 | Vigorous physical activity \|\| id:ebi-a-GCST006098 | Salad / raw vegetable intake \|\| id:ukb-b-1996 | Inverse variance weighted | 20 | 0.31607975 | 0.0657002 | 1.50E-06 |
| **ukb-b-1996** | ebi-a-GCST006098 | Vigorous physical activity \|\| id:ebi-a-GCST006098 | Salad / raw vegetable intake \|\| id:ukb-b-1996 | Simple mode | 20 | 0.39372315 | 0.14841231 | 0.01570282 |
| **ukb-b-1996** | ebi-a-GCST006098 | Vigorous physical activity \|\| id:ebi-a-GCST006098 | Salad / raw vegetable intake \|\| id:ukb-b-1996 | Weighted mode | 20 | 0.39793054 | 0.16695029 | 0.02773826 |
| **ukb-b-6324** | ebi-a-GCST006098 | Vigorous physical activity \|\| id:ebi-a-GCST006098 | Processed meat intake \|\| id:ukb-b-6324 | MR Egger | 23 | 0.13509533 | 0.17451244 | 0.44748219 |
| **ukb-b-6324** | ebi-a-GCST006098 | Vigorous physical activity \|\| id:ebi-a-GCST006098 | Processed meat intake \|\| id:ukb-b-6324 | Weighted median | 23 | -0.0754533 | 0.03693307 | 0.04105493 |
| **ukb-b-6324** | ebi-a-GCST006098 | Vigorous physical activity \|\| id:ebi-a-GCST006098 | Processed meat intake \|\| id:ukb-b-6324 | Inverse variance weighted | 23 | -0.1085666 | 0.03736846 | 0.00366899 |
| **ukb-b-6324** | ebi-a-GCST006098 | Vigorous physical activity \|\| id:ebi-a-GCST006098 | Processed meat intake \|\| id:ukb-b-6324 | Simple mode | 23 | -0.0569462 | 0.08563101 | 0.51295016 |
| **ukb-b-6324** | ebi-a-GCST006098 | Vigorous physical activity \|\| id:ebi-a-GCST006098 | Processed meat intake \|\| id:ukb-b-6324 | Weighted mode | 23 | -0.0357803 | 0.07544894 | 0.64000771 |
| **ukb-b-4616** | ebi-a-GCST006098 | Vigorous physical activity \|\| id:ebi-a-GCST006098 | Nap during day \|\| id:ukb-b-4616 | MR Egger | 89 | -0.1310942 | 0.09139345 | 0.15504488 |
| **ukb-b-4616** | ebi-a-GCST006098 | Vigorous physical activity \|\| id:ebi-a-GCST006098 | Nap during day \|\| id:ukb-b-4616 | Weighted median | 89 | -0.0537114 | 0.0279262 | 0.05443828 |
| **ukb-b-4616** | ebi-a-GCST006098 | Vigorous physical activity \|\| id:ebi-a-GCST006098 | Nap during day \|\| id:ukb-b-4616 | Inverse variance weighted | 89 | -0.0652623 | 0.02490192 | 0.00877309 |
| **ukb-b-4616** | ebi-a-GCST006098 | Vigorous physical activity \|\| id:ebi-a-GCST006098 | Nap during day \|\| id:ukb-b-4616 | Simple mode | 89 | -0.0625663 | 0.0649349 | 0.33792525 |
| **ukb-b-4616** | ebi-a-GCST006098 | Vigorous physical activity \|\| id:ebi-a-GCST006098 | Nap during day \|\| id:ukb-b-4616 | Weighted mode | 89 | -0.0482764 | 0.05502095 | 0.38264819 |
| **ukb-b-3881** | ebi-a-GCST006098 | Vigorous physical activity \|\| id:ebi-a-GCST006098 | Fresh fruit intake \|\| id:ukb-b-3881 | MR Egger | 53 | -0.0277245 | 0.12616196 | 0.82694037 |

| | | | | | | | | |
|---|---|---|---|---|---|---|---|---|
| **ukb-b-3881** | **ebi-a-GCST006098** | **Vigorous physical activity \|\| id:ebi-a-GCST006098** | **Fresh fruit intake \|\| id:ukb-b-3881** | **Weighted median** | **53** | **0.16136177** | **0.0384663** | **2.73E-05** |
| **ukb-b-3881** | ebi-a-GCST006098 | Vigorous physical activity \|\| id:ebi-a-GCST006098 | Fresh fruit intake \|\| id:ukb-b-3881 | Inverse variance weighted | 53 | 0.2103584 | 0.03697867 | 1.28E-08 |
| **ukb-b-3881** | ebi-a-GCST006098 | Vigorous physical activity \|\| id:ebi-a-GCST006098 | Fresh fruit intake \|\| id:ukb-b-3881 | Simple mode | 53 | 0.16760638 | 0.09363947 | 0.07929148 |
| **ukb-b-3881** | ebi-a-GCST006098 | Vigorous physical activity \|\| id:ebi-a-GCST006098 | Fresh fruit intake \|\| id:ukb-b-3881 | Weighted mode | 53 | 0.14738639 | 0.08743063 | 0.09783589 |

**Tests for horizontal pleiotropy**

| id.exposure | id.outcome | outcome | exposure | Egger intercept | se | pval |
|---|---|---|---|---|---|---|
| **ukb-b-1996** | ebi-a-GCST006098 | Vigorous physical activity \|\| id:ebi-a-GCST006098 | Salad / raw vegetable intake \|\| id:ukb-b-1996 | -0.0007564 | 0.00362056 | 0.83686113 |
| **ukb-b-6324** | ebi-a-GCST006098 | Vigorous physical activity \|\| id:ebi-a-GCST006098 | Processed meat intake \|\| id:ukb-b-6324 | -0.0037415 | 0.00262036 | 0.16803755 |
| **ukb-b-4616** | ebi-a-GCST006098 | Vigorous physical activity \|\| id:ebi-a-GCST006098 | Nap during day \|\| id:ukb-b-4616 | 0.00064846 | 0.00086601 | 0.45600425 |
| **ukb-b-3881** | ebi-a-GCST006098 | Vigorous physical activity \|\| id:ebi-a-GCST006098 | Fresh fruit intake \|\| id:ukb-b-3881 | 0.00227091 | 0.00115335 | 0.05440223 |

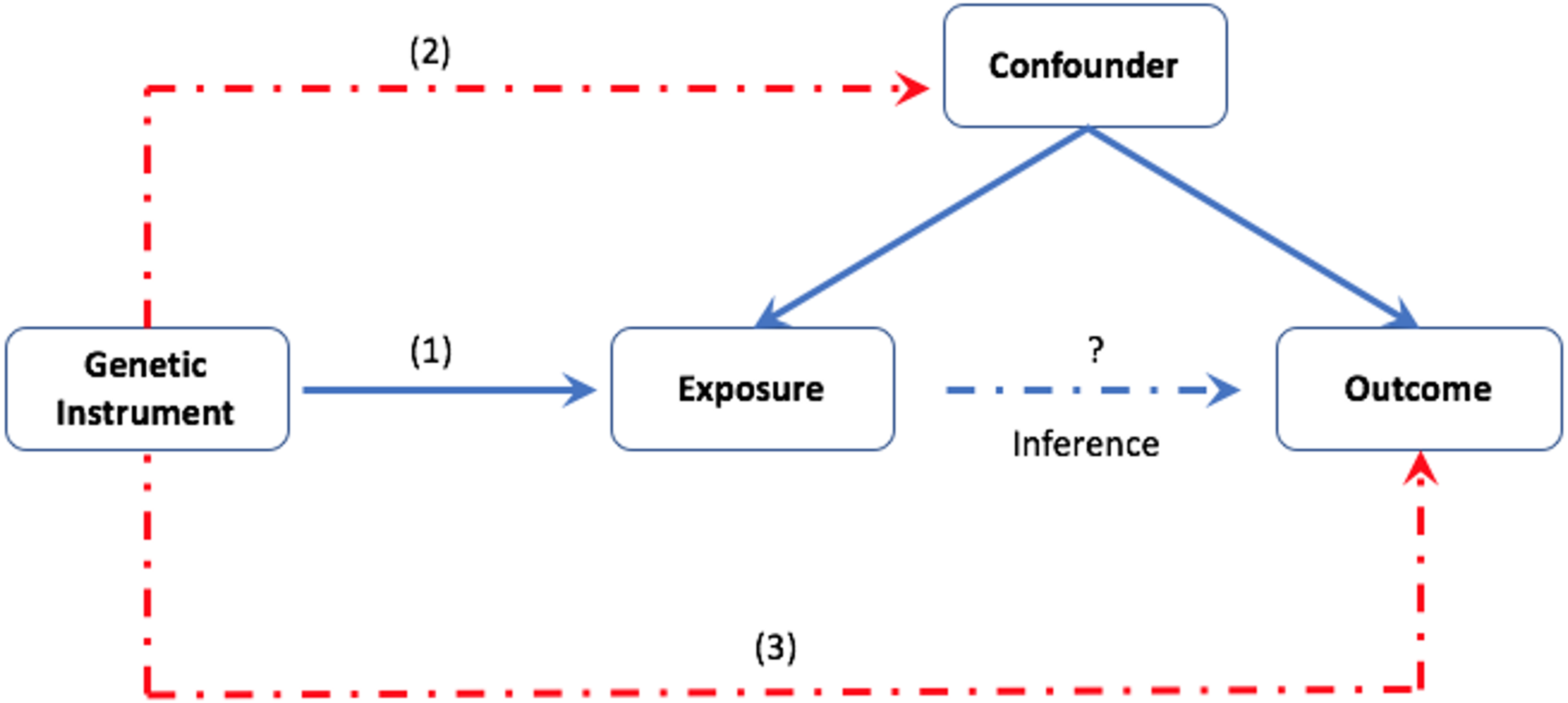


**Figure 1S**. Assumptions of Mendelian randomization: (1) the genetic instrument is associated with the exposure. (2) the genetic instrument should not associate with a confounder. (3) the genetic instrument should affect the outcome only via the exposure.

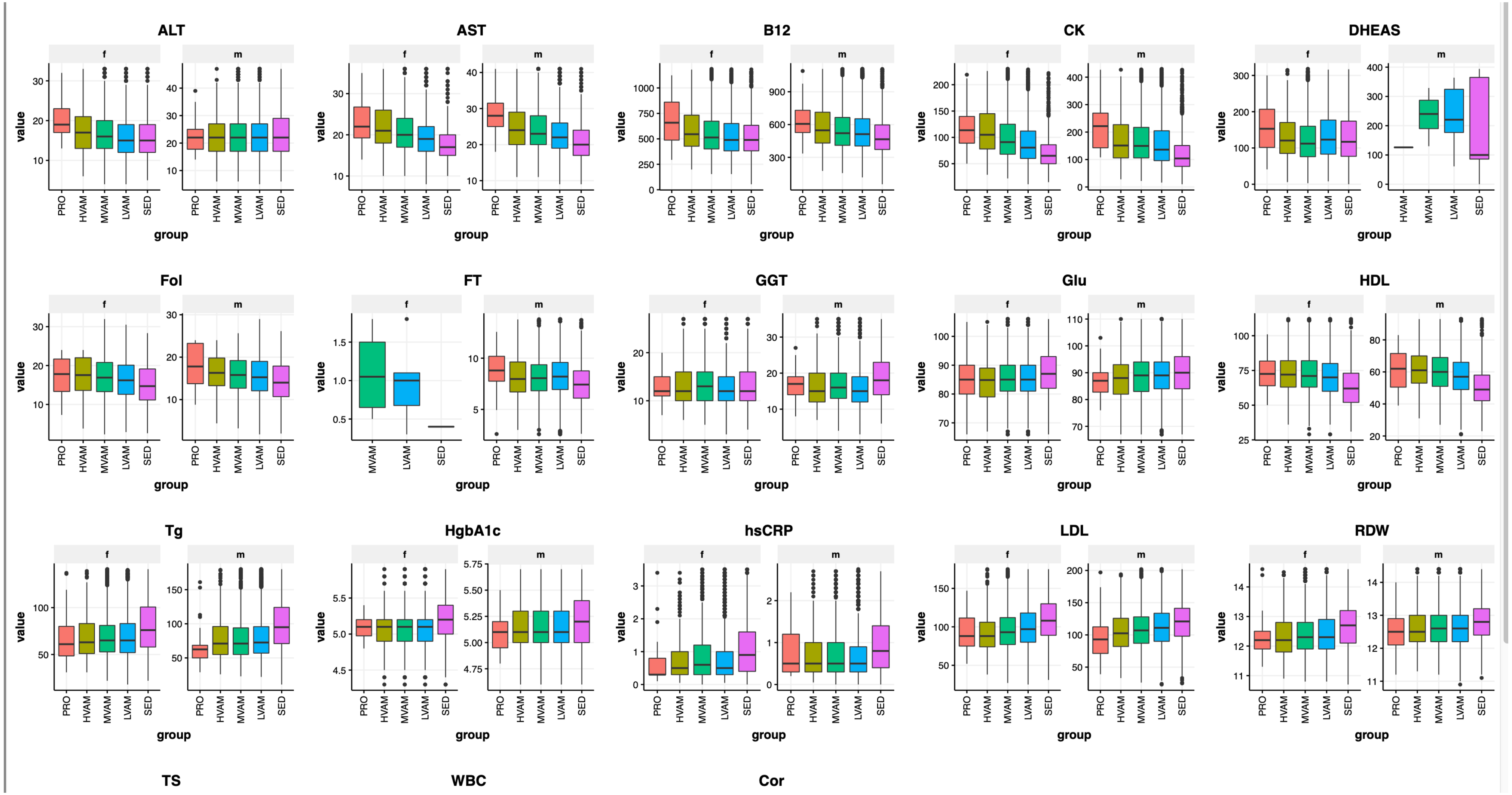


**Figure 2S**. Blood biomarker levels with respect to self-reported running volume and professional athletes in males (m) and females (f).

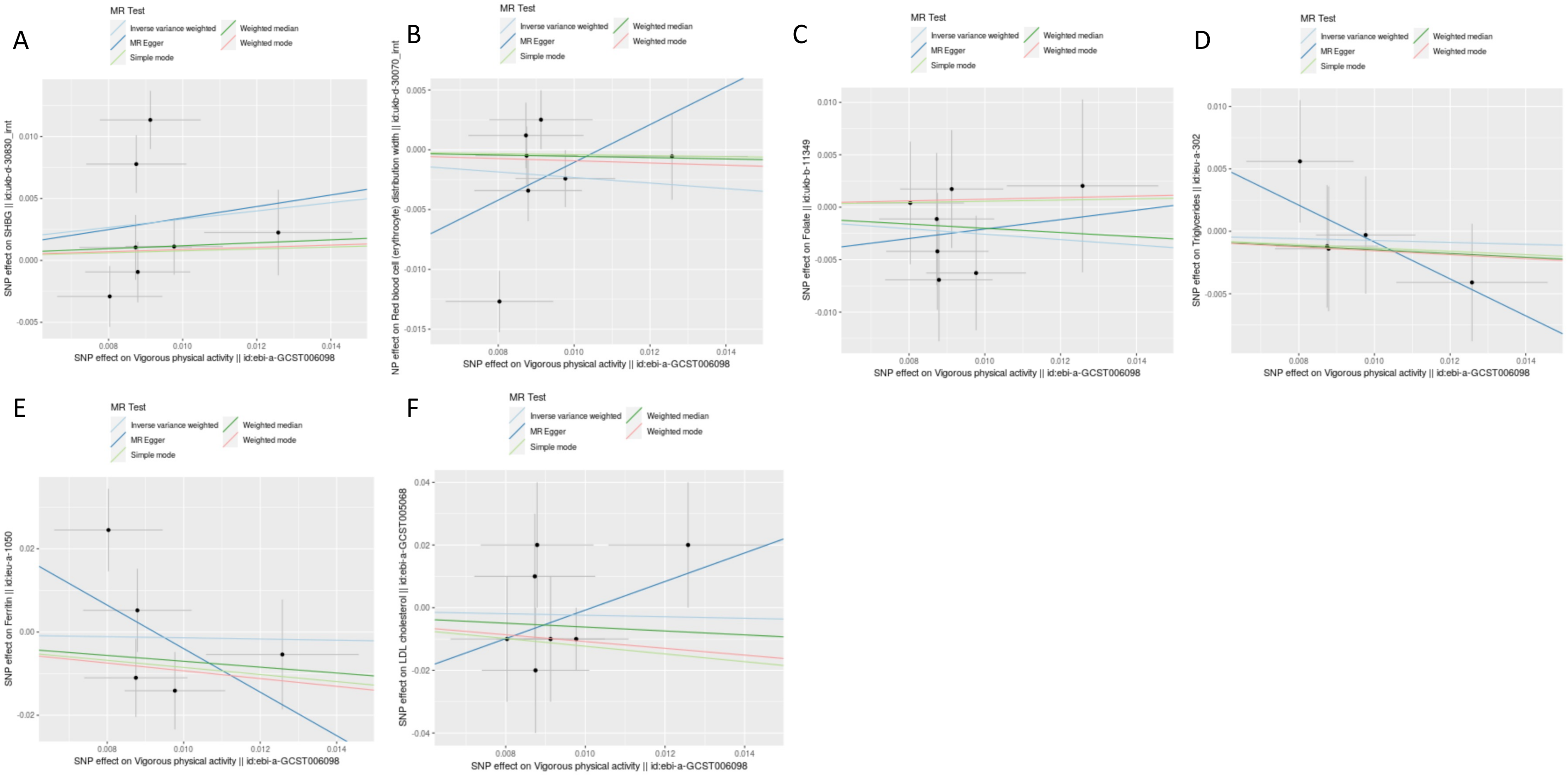


**Figure 3S**. 2S-MR scatter plot showing effects of vigorous physical activity as the exposure on (A) SHBG (B) Red blood cell count (C) Folate (D) Triglycerides (E) Ferritin (F) LDL as outcomes (see Table 5S for statistical significance).

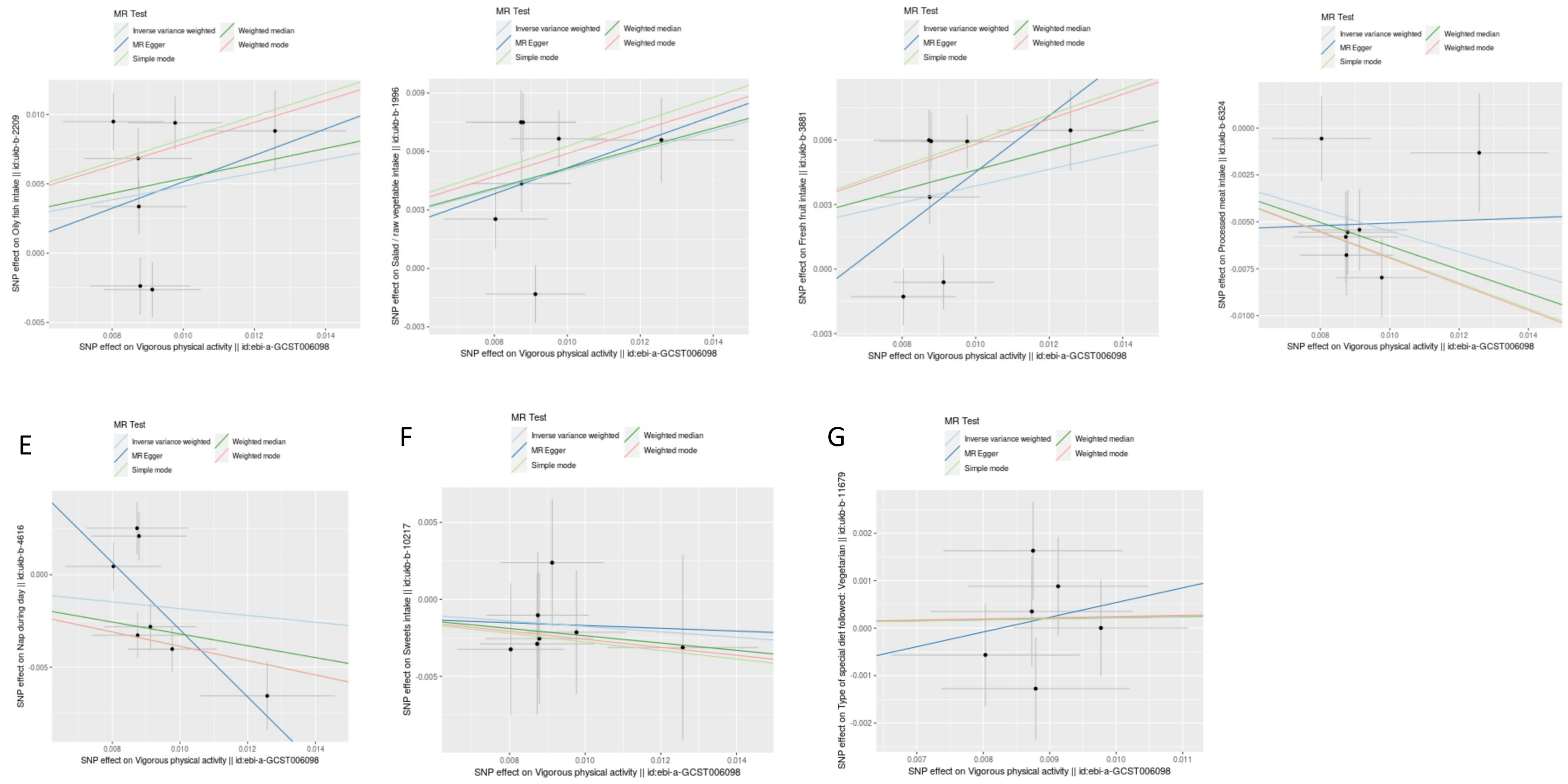


**Figure 4S**. 2S-MR scatter plot showing effects of vigorous physical activity as the exposure on (A) oily fish consumption (B) salad intake(C) fresh fruit intake (D) processed meat intake (E) daytime napping (F) sweets intake (G) vegetarian diet (see Table 6S for statistical significance).

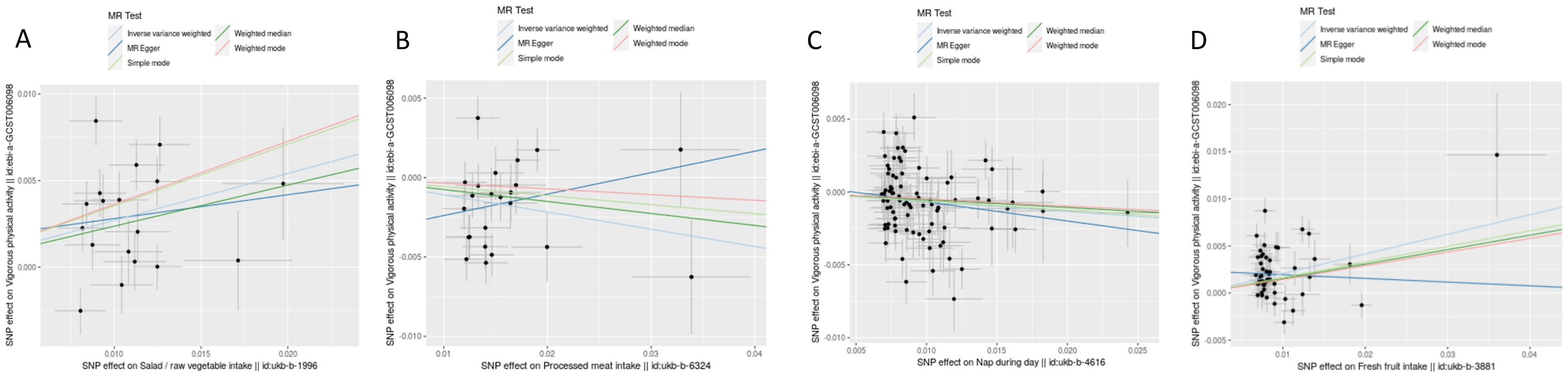


**Figure 5S**. 2S-MR scatter plot showing effects of dietary behaviors as the exposures on vigorous physical activity: (A) salad intake (B) processed meat intake (C) daytime napping (D) fresh fruit intake (see Table 7S for statistical significance).
